# Evolution of COVID-19 Pandemic in India

**DOI:** 10.1101/2020.07.01.20143925

**Authors:** Ali Asad, Siddharth Srivastava, Mahendra K. Verma

## Abstract

A mathematical analysis of patterns for the evolution of COVID-19 cases is key to the development of reliable and robust predictive models potentially leading to efficient and effective governance against COVID-19. Towards this objective, we study and analyze the temporal growth pattern of COVID-19 infection and death counts in various states of India. Our analysis up to August 4, 2020, shows that several states (namely Maharashtra, Tamil Nadu, West Bengal) have reached *t*^2^ power-law growth, while Gujarat and Madhya Pradesh exhibit linear growth. Delhi has reached 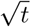 phase and may flatten in the coming days. However, some states have deviated from the universal pattern of the epidemic curve. Uttar Pradesh and Rajasthan show a gradual rise in the power-law regime, which is not the usual trend. Also, Bihar, Karnataka, and Kerala are exhibiting a second wave. In addition, we report that initially, the death counts show similar behavior as the infection counts. Later, however, the death growth rate declines as compared to the infection growth due to better handling of critical cases and increased immunity of the population. These observations indicate that except Delhi, most of the Indian states are far from flattening their epidemic curves.

## 1 Introduction

COVID-19 pandemic has been impacting the life and economy across the globe since December 2019 and has caused major disruptions [37]. As of August 2020, COVID-19 has infected nearly 20 million people across the globe with 90 countries in community transmission stage [38] leading to significant efforts towards control [27], modelling [2,6,12], search for a cure [19] for COVID-19 across the world and India [7,32]. Keeping this in mind, in this paper, we analyze the evolution of COVID-19 cases and deaths in various Indian states. Specifically, we study and model the temporal evolution of infection and death counts for various time intervals and analyze their variations.

At the onset of the COVID-19 pandemic, India imposed the world’s strictest nationwide lockdown beginning from March 25, 2020 [18]. However, preparedness and impact of the lockdown varied across states depending upon past experiences such as the Nipah virus in Kerala or Odisha’s disaster response due to recent natural disasters [18,11]. Therefore, attempts have been made to study the impact of COVID-19 in India. Sardar et al. [28] mathematically assessed the impact of the first 21 days of the lockdown in terms of the total number of cases. Tomar et al. [35] employed deep learning to provide a 30 day forecast of the death cases and recovered cases. Chatterjee et al. [6] provided estimates on the growth of infections using nonpharmacological interventions such as social distancing and lockdown. Network-based epidemic growth models have also been evolved for modeling COVID-19 pandemic [22].

Epidemiological models, e.g. SEIR model, are being evolved to suit the national conditions [4,10,17,20,24]. A model based on delay-differential equations considers the effects of past events [31]. Ranjan [26] studied the effects of various factors in the dynamics of epidemic spread. Due to lack of ample historical data, many models for studying COVID-19 are appearing everyday [9, 3,34,30]. However, none of them is able to model the epidemic pattern to sufficient accuracy [15].

Further, predictive models are based on the underlying patterns of COVID-19 data [25,36]. Note however that the patterns of COVID-19 cases vary due to the extent of government measures [13,14]. Consequently, forecasting COVID-19 is quite complex.

Verma et al. [36] and Chatterjee et al. [8] analyzed infection counts of 21 leading countries. They observed the emergence of power-laws after an initial exponential phase. They showed that China and South Korea followed power-law regimes—*t*^2^, *t*, 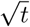— before flattening their epidemic curves. Also, the infection data for European countries (Spain, France, Italy, and Germany), USA, and Japan followed a power-law regime (*t^n^*, 1 ≤ *n* ≤ 4). They attributed these characteristics to long-distance travel and asymptomatic carriers. They concluded that 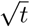 regime is a common feature among all infection curves that exhibit saturation.

In this paper, we extend the works of Verma et al. [36] and Chatterjee et al. [8] to the severely affected Indian states. We observe that some states exhibit *t*^2^ and *t* growth phases, while some others have linear or 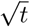 growths. Bihar, Karnataka, and Kerala appear to have second waves of infections. These findings will be useful to the epidemic control panel. We discuss our results in Section 2 and conclude in Section 3

## 2 Analysis and Results

In this paper, we analyze the COVID-19 infection and death counts in nineteen Indian states: Maharashtra, Tamil Nadu, Delhi, Gujarat, Uttar Pradesh, Rajasthan, Madhya Pradesh, West Bengal, Karnataka, Bihar, and Kerela.We combine the data of all the north-eastern (NE) states (Arunachal Pradesh, Assam, Manipur, Meghalaya, Mizoram, Nagaland, Sikkim, and Tripura) because the counts for each of them is rather small for any statistical analysis. As of August 4, 2020, the above states constitute about 78% (1.45×10^6^/=1.85×10^6^) of the total COVID-19 infections in India. For our analysis, we employed the real-time data available at the website of *Ministry of Health and Family Welfare, Government of India* [23]. We have consolidated the data using the Application Programming Interface (APIs) from COVID-19 India Tracker [1].

For our analysis, we consider data till August 4, 2020. First, we perform a temporal evolution analysis of Infection count, which is denoted by *I*(*t*), where *t* is time in days. In Fig. 1, we plot the time series of *I*(*t*) and its derivative *İ*(*t*) in semi-logy (the *y*-axis has logarithmic scale, while the *x*-axis has linear scale) format using red and blue curves respectively. The starting date, listed in Table 1, is chosen from the day the infection began to increase in the respective states.

**Fig. 1.**
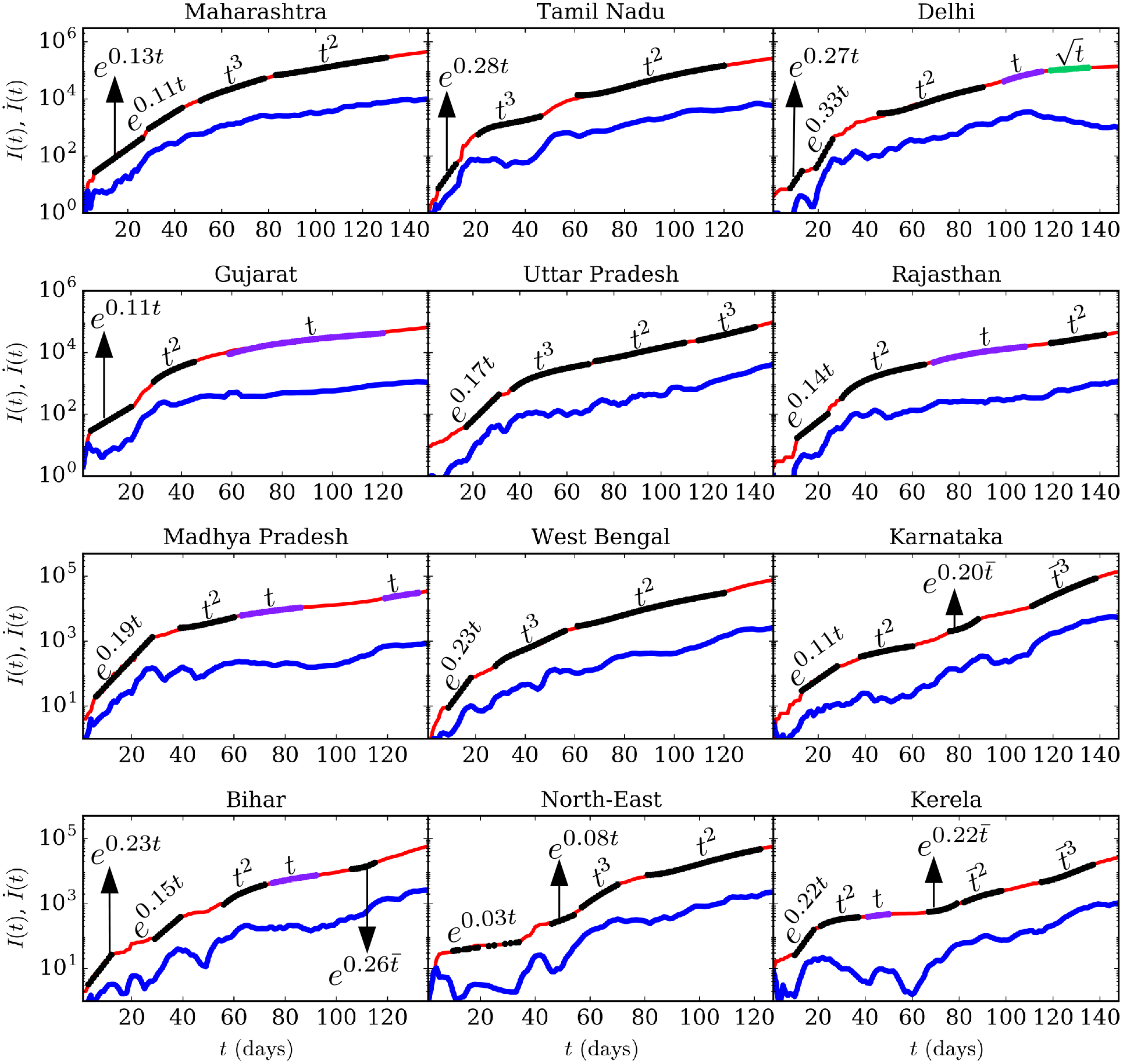
(color online) The *semi-logy* plots of total infection count (*I*(*t*)) vs. time (*t*) (red thin curves) and *İ*(*t*) vs. *t* (blue thick curves) for the eleven states individually and consolidated for north-eastern Indian states. The dotted curves represent the best-fit curves. Refer Table 1 for the best-fit functions.

**Table 1.** Best-fit functions for the total infections and the corresponding relative errors for major Indian states. The order of the functions for respective states correspond to the best-fit curves marked on *I*(*t*) of Fig 1.

| States<br>(Start Date) | Best-fit functions with errors |
| --- | --- |
| Maharashtra<br>(March 10) | 1) $12e^{0.13t}$ ($\pm 6.9\%$)<br>2) $30e^{0.11t}$ ($\pm 5.2\%$)<br>3) $0.9t^3 - 140t^2 + 7900t - 15 \times 10^4$ ($\pm 1.0\%$)<br>4) $70t^2 - 9700t + 41 \times 10^4$ ($\pm 1.2\%$) |
| Tamil Nadu<br>(March 18) | 1) $1.9e^{0.28t}$ ($\pm 7.3\%$)<br>2) $0.15t^3 - 15t^2 + 530t - 5400$ ($\pm 2.0\%$)<br>3) $41t^2 - 5100t + 17 \times 10^4$ ($\pm 4.1\%$) |
| Delhi<br>(March 10) | 1) $0.8e^{0.27t}$ ($\pm 5.7\%$)<br>2) $0.07e^{0.33t}$ ($\pm 10.0\%$)<br>3) $13t^2 - 1200t + 32000$ ($\pm 7.0\%$)<br>4) $3100t - 27 \times 10^4$ ($\pm 1.2\%$)<br>5) $37000\sqrt{t} - 30 \times 10^4$ ($\pm 0.48\%$) |
| Gujarat<br>(March 20) | 1) $19e^{0.11t}$ ($\pm 4.3\%$)<br>2) $3t^2 + 9t - 1700$ ($\pm 4.3\%$)<br>3) $550t - 23000$ ($\pm 4.7\%$) |
| Uttar Pradesh<br>(March 10) | 1) $2.1e^{0.17t}$ ($\pm 6.8\%$)<br>2) $0.01t^3 - 1.7t^2 + 180t - 4400$ ($\pm 1.5\%$)<br>3) $7.5t^2 - 960t + 36000$ ($\pm 1.8\%$)<br>4) $0.7t^3 - 230t^2 + 26000t - 98 \times 10^4$ ($\pm 0.27\%$) |
| Rajasthan<br>(March 10) | 1) $3.8e^{0.14t}$ ($\pm 7.0\%$)<br>2) $0.55t^2 + 54t - 1800$ ($\pm 2.6\%$)<br>3) $270t - 14000$ ($\pm 1.4\%$)<br>4) $13t^2 - 2700t + 15 \times 10^4$ ($\pm 0.45\%$) |
| Madhya Pradesh<br>(March 21) | 1) $6.3e^{0.19t}$ ($\pm 9.7\%$)<br>2) $4.7t^2 - 330t + 8300$ ($\pm 1.6\%$)<br>3) $200t - 6400$ ($\pm 0.38\%$)<br>4) $750t - 68000$ ($\pm 0.24\%$) |
| West Bengal<br>(March 18) | 1) $1.1e^{0.23t}$ ($\pm 7.2\%$)<br>2) $0.1t^3 - 9t^2 + 330t - 3900$ ($\pm 4.8\%$)<br>3) $7t^2 - 790t + 25000$ ($\pm 4.9\%$) |
| Karnataka<br>(March 10) | 1) $6.7e^{0.11t}$ ($\pm 6.5\%$)<br>2) $0.1t^2 + 9.7t - 140$ ($\pm 1.2\%$)<br>3) $230e^{0.20\bar{t}}$ ($\pm 7.8\%$)<br>2) $1.03\bar{t}^3 - 75\bar{t}^2 + 2400\bar{t} - 28000$ ($\pm 0.8\%$) |
| Bihar<br>(March 22) | 1) $1.6e^{0.23t}$ ($\pm 9.0\%$)<br>2) $0.9e^{0.15t}$ ($\pm 3.2\%$)<br>3) $0.87t^2 + 69t - 5700$ ($\pm 2.9\%$)<br>4) $185t - 9500$ ($\pm 1.05\%$)<br>5) $580e^{0.25\bar{t}}$ ($\pm 4.3\%$) |
| Kerela<br>(March 10) | 1) $2.8e^{0.22t}$ ($\pm 8.0\%$)<br>2) $0.02t^3 - 2.3t^2 + 92t - 900$ ($\pm 1.0\%$)<br>3) $9.3t + 14$ ($\pm 0.81\%$)<br>4) $26e^{0.22\bar{t}}$ ($\pm 6.8\%$)<br>5) $0.1\bar{t}^2 + 83\bar{t} - 780$ ($\pm 2.3\%$)<br>6) $0.18\bar{t}^3 - 12\bar{t}^2 - 15\bar{t} + 12000$ ($\pm 0.64\%$) |
| North East<br>(March 10) | 1) $28e^{0.03t}$ ($\pm 6.6\%$)<br>2) $7e^{0.08t}$ ($\pm 4.1\%$)<br>3) $0.14t^3 - 16t^2 + 600t - 6400$ ($\pm 2.1\%$)<br>4) $23t^2 - 3600t - 15000$ ($\pm 2.0\%$) |

Similarly, we studied the evolution of total death cases for six states (Maharashtra, Delhi, Gujarat, Tamil Nadu, West Bengal, and Uttar Pradesh) that have reported a large number of deaths. The cumulative death cases are denoted by *D*(*t*). The time series of *D*(*t*) and its derivative 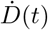 are plotted in Fig. 2 in semi-logy using red and blue curves respectively. Also, the starting date (see Table 2) is considered from the day death counts begin to increase. The starting dates for the infection plots and the death plots are not the same. This is because the death cases peaks after a delay from the infection peak due to the incubation period.

**Fig. 2.**
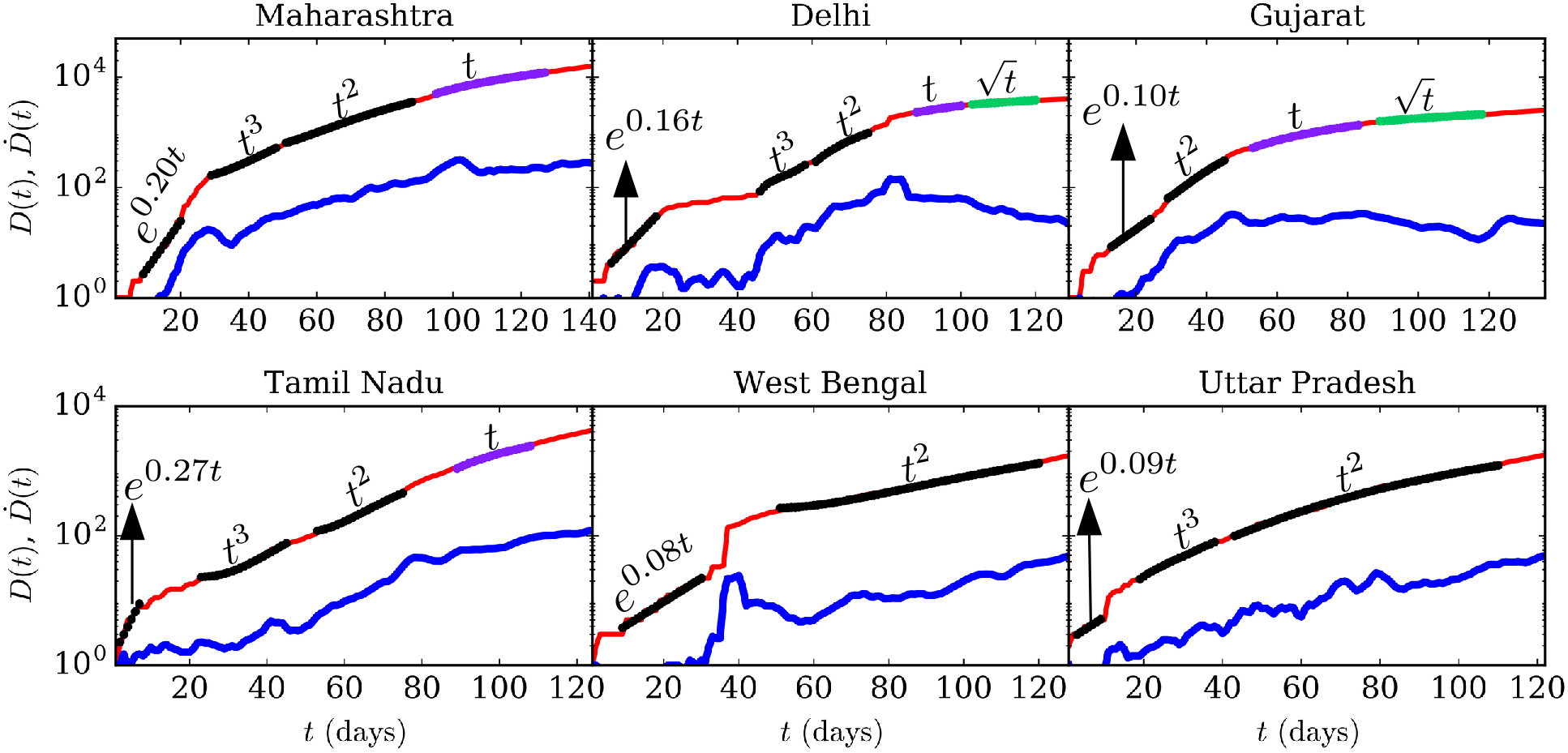
(color online) The *semi-logy* plots of total death cases (*D*(*t*)) vs. time (*t*) (red thin curves) and 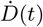 vs. *t* (blue thick curves) for six states of India. The dotted curves represent the best-fit curves (see Table 2).

**Table 2.** Best-fit functions for the death cases and corresponding relative errors for major Indian states. The order of the functions for respective states correspond to the best-fit curves on *D*(*t*) of Fig 2.

| States<br>(Start Date) | Best-fit functions with errors |
| --- | --- |
| Maharashtra<br>(March 17) | 1) $0.44e^{0.20t}$ ($\pm 6.4\%$)<br>2) $0.02t^3 - 1.1t^2 + 30t - 170$ ($\pm 1.1\%$)<br>3) $1.4t^2 - 120t + 3000$ ($\pm 1.3\%$)<br>4) $220t - 16000$ ($\pm 1.5\%$) |
| Delhi<br>(March 29) | 1) $1.6e^{0.16t}$ ($\pm 9.6\%$)<br>2) $0.13t^3 - 20t^2 + 1000t - 17000$ ($\pm 2.9\%$)<br>3) $0.57t^2 - 30t - 22$ ($\pm 1.6\%$)<br>4) $60t - 3200$ ($\pm 0.13\%$)<br>5) $740\sqrt{t} - 4200$ ($\pm 0.27\%$) |
| Gujarat<br>(March 22) | 1) $2.2e^{0.10t}$ ($\pm 4.1\%$)<br>2) $0.6t^2 - 29t + 400$ ($\pm 3.3\%$)<br>3) $28t - 970$ ($\pm 0.98\%$)<br>4) $370\sqrt{t} - 1900$ ($\pm 0.63\%$) |
| Tamil Nadu<br>(April 03) | 1) $1.3e^{0.27t}$ ($\pm 8.9\%$)<br>2) $0.002t^3 - 0.01t^2 + 0.96t + 25$ ($\pm 2.9\%$)<br>3) $0.57t^2 - 58t + 1600$ ($\pm 2.3\%$)<br>4) $70t - 5100$ ($\pm 1.5\%$) |
| West Bengal<br>(March 30) | 1) $1.8e^{0.08t}$ ($\pm 8.6\%$)<br>2) $0.20t^2 - 20t + 740$ ($\pm 3.2\%$) |
| Uttar Pradesh<br>(April 04) | 1) $2.27e^{0.1t}$ ($\pm 5.5\%$)<br>2) $0.003t^3 - 0.2t^2 + 6.2t - 48$ ($\pm 2.6\%$)<br>3) $0.17t^2 - 10t + 200$ ($\pm 3.4\%$) |

We employ exponential and polynomial functions to compute best-fit curves on different regions of *I*(*t*) and *D*(*t*) data. The time series for both *I*(*t*) and *D*(*t*) follow exponential regimes during the early phases of the pandemic and subsequently transition to power-law regimes. This is in accordance with the earlier work of Verma et al. [36] and Chatterjee et al. [8]. The bestfit functions along with their relative errors are listed in Tables 1 and 2 for the infected and death cases respectively. The error for a given fit is calculated using the mean of the absolute difference between the best-fit curve and the corresponding actual curve. In Table 1, we report *rel*. *error* = (| *I_fit_ − I_actual_* | ×100)/*I_actual_*.

The epidemic curves transition to power-law regimes after the exponential phase. We employ Python’s *polyfit* function to calculate the best-fit polynomials for these regions. The function *polyfit* employs regression via minimization of error and provides best-fit curves. Verma et al. [36] and Chatterjee et al. [8] showed that the epidemic curves of many countries pass through a series of polynomials, *t*^3^, *t*^2^, *t*, 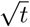, before saturation. The intermediate power-law regimes are believed to appear due to lockdown and social restrictions. As shown in Figure 1, many states deviate from the above patterns, which are possibly due to unlocking in India on June 8. Note that the unlocking of various states occurred at a later date. In the following discussion, we describe how the *I*(*t*) curves for various states behave.

The infection curves of Maharashtra, Tamil Nadu, and West Bengal, as well as the combined NE-states, exhibit a *t*^3^ regime followed by a *t*^2^ phase. Whereas, Gujarat and Madhya Pradesh have reached a linear growth after going through a *t*^2^ regime. The *I*(*t*) curve of Delhi follows *t*^2^ and linear regimes before reaching a 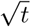 growth. This trend indicates that Delhi is close to saturating its epidemic curve. It is worth noting that the states mentioned above follow the universal pattern of the epidemic curve [8].

Unfortunately, the power-law regime of some states does not follow the universal trend. For instance, Uttar Pradesh reached a *t*^3^ phase after passing through *t*^3^ and *t*^2^ regimes. Also, the infection curve of Rajasthan reached *t*^2^ after following *t*^2^ and linear regimes. These states observed a gradual growth in daily cases as their *I*(*t*) curves pass through the power-law regime. However, this growth is still not exponential and hence does not amount to a second wave. Note that such deviations from the universal pattern are indicators for authorities to take suitable action.

The infection curves of Bihar, Kerela, and Karnataka exhibit a rise which is preceded by a region of a linear regime or a nearly flattened curve. Also, this growth of the *I*(*t*) curve is exponential indicating a second wave for the epidemic [5]. The emergence of this phase corresponds with relaxation in lockdowns and an increase in testing intensity. In Bihar, such a surge may have resulted from the influx of migrant workers and students from different parts of the country. The best-fit curves for the second wave are functions of 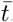, where 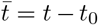. Here, *t*_0_ corresponds to the day from which the daily count shows an unprecedented rise after a region of decline.

Similar to the *I*(*t*) curves, the *D*(*t*) curves begin with exponential regimes (*D*(*t*) = *A_d_* exp(*β_d_*(*t*))), and then transition to power-law regimes (*t*^3^, *t*^2^, *t*). Interestingly, for many states, the power-laws for both *I*(*t*) and *D*(*t*) curves are qualitatively similar. For example, both *I*(*t*) and *D*(*t*) curves for Gujarat exhibit a *t*^2^ region followed by a linear phase (*t*). This further substantiates the claims of Chatterjee et al. [8] that *D*(*t*) is proportional to *I*(*t*) statistically. This is because a fraction of the infected population is susceptible to death. Note that the growth of *D*(*t*) curve has declined in many states with respect to their *I*(*t*) curves. This may be attributed to immunity developed in the community, better handling of critical cases, plasma therapy, etc.

The values of *β_i_* and *β_d_* represent the growth rates of infected and death cases, respectively. It must be noted that *β_i_* and *β_d_* depend on various factors such as immunity level, the average age of the population, population density, local policy decisions (lockdowns, testing intensity, social distancing, healthcare facilities), etc.

In Figs. 1 and 2, we also plot daily infection and death counts, which are represented by *İ*(*t*) and 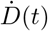 respectively. We calculate the derivative using Python’s *gradient* function and take a 5-day moving average in order to smoothen the *İ*(*t*) and 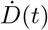 curves. We observe that in the exponential regimes, the daily counts are proportional to the cumulative number of infected and death cases i.e. *İ* ≈ *β_i_I* and 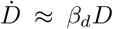. Verma et al. [36] show that power-law regime can be approximated as *I*(*t*) ~ *At^n^*, and hence, *İ* ~ *I*^1−1/^*^n^*. Similarly, it can be shown that for power-laws 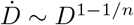. This shows that the daily counts are suppressed in the power-law region compared to the exponential phase. Note that in the linear growth regime, 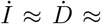 constant, implying a constant daily count. The daily count is expected to decrease after a linear regime (see Delhi in Fig. 1), however, this may not be the case when a second wave emerges.

An interesting question is whether the Indian states with lower COVID-19 cases are closer to saturation. To investigate this issue, we compute the infection time series for India without the three worst affected states, which are Maharashtra, Tamil Nadu, and Delhi. We denote this time series as *Ī*/(*t*), and it is computed as *Ī*(*t*) = *I*(*t*)*_IND_* − {*I*(*t*)*_MH_* +*I*(*t*)*_TN_* +*I*(*t*)*_DL_*}. In Fig. 3, we plot *Ī*(*t*) and 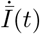, and compare them with the total *I*(*t*) and *İ*(*t*). From the plots it is evident that both the plots exhibit exponential and power-law regimes (see Table 3), and that *Ī*(*t*) and *I*(*t*) are proportional to each other. Although these states comprise of almost 45% (85 × 10^6^/185 × 10^6^) of the total Infection count in India, their removal from total *I*(*t*) does not cause any behavioural change in the *Ī*(*t*) curve. Based on these observations we conclude that almost all the affected states shown in Fig. 1 are following similar epidemic evolution.

**Fig. 3.**
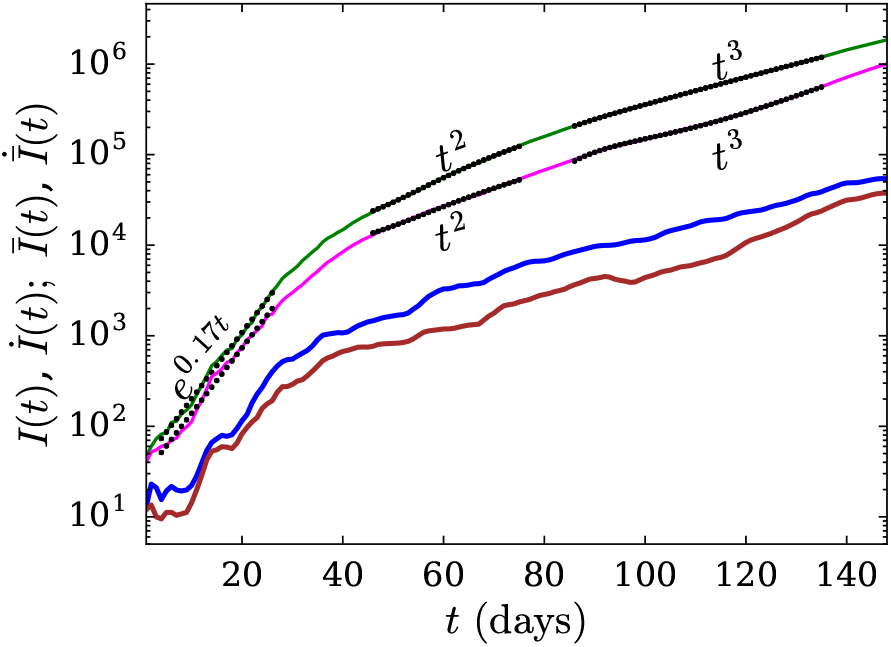
(color online) The *semi-logy* plot of total Infection count (*I*(*t*)) vs. time (*t*) curves for India (green curve) and India other than Maharashtra, Tamil Nadu and Delhi (magenta curve) where, *Ī*(*t*) = *I*(*t*)*_IND_* − {*I*(*t*)*_MH_* +*I*(*t*)*_TN_* +*I*(*t*)*_DL_*}. The thick blue and brown curves in the plot depict the derivatives of *I*(*t*) and *Ī*(*t*) respectively. The dotted curves represent the best-fit curves.

**Table 3.**
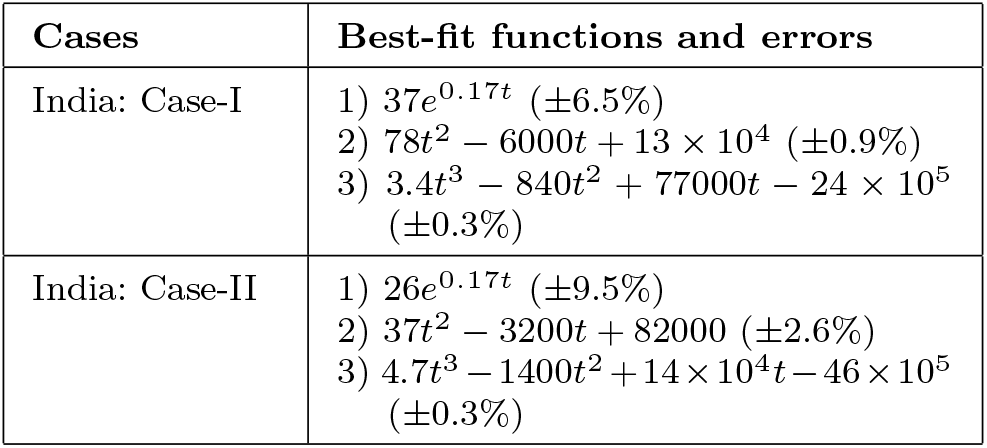
The best-fit functions for cumulative cases and the respective relative errors for various stages of evolution shown in Fig. 3.

In Fig. 4, we plot *I*(*t*) vs. *t* curve in log-log (both *x*-axis and *y*-axis has a logarithmic scale) format for both cases shown in Fig. 3. In the power-law region, we fit a power-law (*I*(*t*) = *At^n^*) instead of a polynomial curve. The exponential *n* of power-law fit (*n* = 4) differs from that calculated using polynomial fit (*n* = 2, 3). This analysis indicates that for a epidemic curve, the power-law exponent is typically larger than the highest power of the corresponding polynomials (the best-fit curve). Still, higher-order polynomials will lead to larger power-law exponent.

**Fig. 4.**
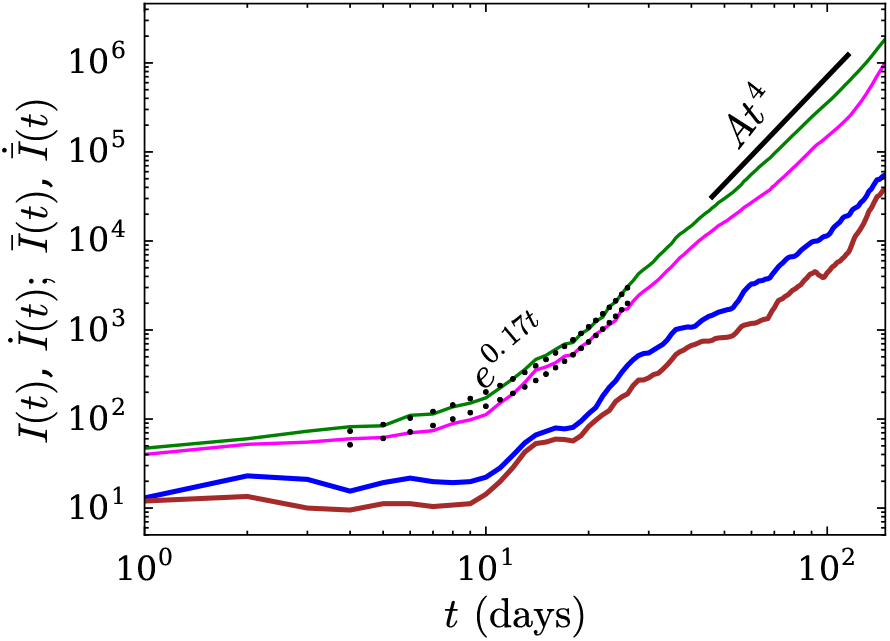
(color online) The *log-log* plot of total infected individual vs. time for India (both cases, see Table 3). Both *I*(*t*) (green curve) and *Ī*(*t*) (magenta curve) curves follow a power-law, i.e., *I*(*t*) = *At*^4^, where *A* = 0.007. The thick blue and brown curves in the plot depict the derivatives of *I*(*t*) and *Ī*(*t*) respectively.

We can summarize the findings of the state-wise epidemic study as follows. Most of the Indian states exhibit rise in the growth of infected cases. Some have reached up to *t*^2^ part of the epidemic evolution, while others have reached the linear regime (*I*(*t*) ~ *t*). Unfortunately, Uttar Pradesh and Rajasthan show an increasing trend in the power-law phase. While Bihar, Kerala, and Karnataka are observing a second wave of the epidemic. However, Delhi exhibits a decrease in daily cases and is closer to saturation. The overall count in India has shifted from *t*^2^ to *t*^3^. These observations indicate that we are far from saturation or flattening of the epidemic curve.

We conclude in the next section.

## 3 Discussions and Conclusions

In this paper, we analyzed the cumulative infection and death counts of the COVID-19 epidemic in the worst-affected states of India. The respective time series, *I*(*t*) and *D*(*t*), exhibit exponential and power-law growth in the epidemic. Maharashtra, Tamil Nadu, and West Bengal and combined NE-states have reached *t*^2^ growth. While Gujarat and Madhya Pradesh have reached linear phase. The infection rate in Delhi exhibits a 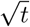 regime which indicates that it is close to flattening its curve. All these states follow the universal trend of the epidemic curve. However, Uttar Pradesh and Rajasthan, as well as states exhibiting a second wave (Bihar, Kerela, and Karnataka) deviate from the universal pattern. We remark that such deviations are indicators for the authorities to take suitable action. The epidemic in India has grown alarmingly after the lockdown restrictions were lifted. Note that the lifting of lockdown is expected to increase the social contacts, and hence the epidemic growth.

Regarding the death count, among the six states we analysed, West Bengal and Uttar Pradesh exhibit *t*^2^ growth, while Tamil Nadu and Maharashtra show linear growth. Delhi and Gujarat have reached 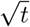 regime. These observations indicate that the death rate exhibits a decline as compared to the growth rate of the infected cases. This may be attributed to immunity developed in the population [33,16] and better treatment of critical cases (plasma therapy, more ventilators, early detection, etc.). At the initial stage, the death rate and infection rate are nearly proportional to each other, consistent with the earlier observation of Chatterjee et al. [8]. We also observe that at present, the infection count in the whole country is increasing as *t*^3^. These observations indicate that we are far from the flattening of the epidemic curve.

The present work is based on data analytics, rather than focussing on specific epidemic models which are being constantly revised in order to successfully forecast the epidemic evolution [24,20,21]. Note, however, that the epidemic models involve many free parameters that lead to ambiguities and difficulties in the forecasting of the epidemic evolution. Our focus on data analytics is due to the latter reason. Our work shows that the power laws in the epidemic curves indicate the stage of the epidemic evolution. This feature helps us in contrasting the evolution of the COVID-19 epidemic in various states of India.

## Data Availability

All data used is available online

## Acknowledgements

The authors thank Soumyadeep Chatterjee and Shashwat Bhattacharya for their help in early works. We also thank Shayak Bhattacharya, Prateek Sharma, and Anurag Gupta for useful discussions. This project is supported by a SERB MATRICS project SERB/F/847/2020-2021. Ali Asad is supported by Indo-French (CEFIPRA) project 6104-1.

## References

1. Covid19 india tracker. URL https://coronaindia.github.io

2. Barkur, G., Vibha, G.B.K.: Sentiment analysis of nationwide lockdown due to covid 19 outbreak: Evidence from india. Asian journal of psychiatry (2020)

3. Bhardwaj, R.: A predictive model for the evolution of covid-19. Transactions of the Indian National Academy of Engineering (2020). DOI10.1007/s41403-020-00130-w. URL https://doi.org/10.1007/s41403-020-00130-w

4. Bjørnstad, O.N.: Epidemics: Models and Data using R. Springer (2018)

5. de Castro, F.: Modelling of the second (and subsequent) waves of the coronavirus epidemic. spain and germany as case studies. medRxiv (2020). DOI 10.1101/2020.06.12.20129429. URL https://www.medrxiv.org/content/early/2020/07/28/2020.06.12.20129429

6. Chatterjee, K., Chatterjee, K., Kumar, A., Shankar, S.: Healthcare impact of covid-19 epidemic in india: A stochastic mathematical model. Medical Journal Armed Forces India (2020)

7. Chatterjee, P., Nagi, N., Agarwal, A., Das, B., Banerjee, S., Sarkar, S., Gupta, N., Gangakhedkar, R.R., et al.: The 2019 novel coronavirus disease (covid-19) pandemic: A review of the current evidence. Indian Journal of Medical Research 151(2), 147 (2020)

8. Chatterjee, S., Asad, A., Shayak, B., Bhattacharya, S., Alam, S., Verma, M.K.: Evolution of covid-19 pandemic: Power-law growth and saturation. Journal of Indian Statistical Association 58(1), 1–31 (2020). URL https://sites.google.com/site/indianstatisticalassociation/journal/journalprevious-volumes/june-2020

9. Chauhan, P., Kumar, A., Jamdagni, P.: Regression analysis of covid-19 spread in india and its different states. medRxiv (2020). DOI 10.1101/2020.05.29.20117069. URL https://www.medrxiv.org/content/early/2020/05/29/2020.05.29.20117069

10. Daley, D.J., Gani, J.: Epidemic Modelling: An Introduction. Cambridge University Press (2001)

11. Dore, B.: Covid-19: collateral damage of lockdown in india. BMJ 369 (2020)

12. Giordano, G., Blanchini, F., Bruno, R., Colaneri, P., Di Filippo, A., Di Matteo, A., Colaneri, M.: Modelling the covid-19 epidemic and implementation of population-wide interventions in italy. Nature Medicine pp. 1–6 (2020)

13. Hale, T., Petherick, A., Phillips, T., Webster, S.: Variation in government responses to covid-19. Blavatnik school of government working paper 31 (2020)

14. Hale, T., Webster, S., Petherick, A., Phillips, T., Kira, B.: Oxford covid-19 government response tracker. Blavatnik School of Government 25 (2020)

15. Holmdahl, I., Buckee, C.: Wrong but usefulwhat covid-19 epidemiologic models can and cannot tell us. New England Journal of Medicine (2020)

16. Kwok, K.O., Lai, F., Wei, W.I., Wong, S.Y.S., Tang, J.W.: Herd immunity–estimating the level required to halt the covid-19 epidemics in affected countries. Journal of Infection 80(6), e32–e33 (2020)

17. Labadin, J., Hong, B.H.: Transmission Dynamics of 2019-nCoV in Malaysia. medrxiv.org (doi:0.1101/2020.02.07.20021188) (2020)

18. Lancet, T.: India under covid-19 lockdown. Lancet (London, England) 395(10233), 1315 (2020)

19. Le, T.T., Andreadakis, Z., Kumar, A., Roman, R.G., Tollefsen, S., Saville, M., Mayhew, S.: The covid-19 vaccine development landscape. Nat Rev Drug Discov 19(5), 305–306 (2020)

20. López, L.R., Rodo, X.: A modified SEIR model to predict the COVID-19 outbreak in Spain and Italy: simulating control scenarios and multi-scale epidemics. medarxiv.org (doi:10.1101/2020.03.27.20045005) (2020)

21. Mandal, S., Bhatnagar, T., Arinaminpathy, N., Agarwal, A., Chowdhury, A., Murhekar, M., Gangakhedkar, R., Sarkar, S.: Prudent public health intervention strategies to control the coronavirus disease 2019 transmission in India: A mathematical model-based approach. Indian Journal of Medical Research (preprint) (2020)

22. Marathe, M., Vullikanti, A.K.S.: Computational epidemiology. Commun. ACM 56(7), 88–96 (2013)

23. Ministry of Health and Family Welfare, Govt. of India: URL https://www.mohfw.gov.in/

24. Peng, L., Yang, W., Zhang, D., Zhuge, C., Hong, L.: Epidemic analysis of COVID-19 in China by dynamical modeling. medarxiv.org (doi:10.1101/2020.03.14.20036202) (2020)

25. Petropoulos, F., Makridakis, S.: Forecasting the novel coronavirus covid-19. PloS one 15(3), e0231236 (2020)

26. Ranjan, R.: Temporal Dynamics of COVID-19 Outbreak and Future Projections: A Data-Driven Approach. Transactions of the Indian National Academy of Engineering (2020). DOI10.1007/s41403-020-00112-y. URL https://doi.org/10.1007/s41403-020-00112-y

27. Rawaf, S., Yamamoto, H.Q., Rawaf, D.: Unlocking towns and cities: Covid-19 exit strategy. East Mediterr Health J 26(5), 499–502 (2020)

28. Sardar, T., Nadim, S.S., Chattopadhyay, J.: Assessment of 21 days lockdown effect in some states and overall india: a predictive mathematical study on covid-19 outbreak. arXiv preprint arXiv:2004.03487 (2020)

29. Schüttler, J., Schlickeiser, R., Schlickeiser, F., Kroger, M.: Covid-19 Predictions Using a Gauss Model, Based on Data from April 2. preprints.org (2020)

30. Sharma, V.K., Nigam, U.: Modeling and forecasting for covid-19 growth curve in india. medRxiv (2020). DOI 10.1101/2020.05.20.20107540. URL https://www.medrxiv.org/content/early/2020/05/28/2020.05.20.20107540

31. Shayak, B., Rand, R.H.: Self-burnout - a new path to the end of covid-19. medrxiv.org (doi:10.1101/2020.04.17.20069443) (2020)

32. Singhal, T.: A review of coronavirus disease-2019 (covid-19). The Indian Journal of Pediatrics pp. 1–6 (2020)

33. Tay, M.Z., Poh, C.M., Rénia, L., MacAry, P.A., Ng, L.F.: The trinity of covid-19: immunity, inflammation and intervention. Nature Reviews Immunology pp. 1–12 (2020)

34. Tiwari, A.: Modelling and analysis of covid-19 epidemic in india. medRxiv (2020). DOI 10.1101/2020.04.12.20062794. URL https://www.medrxiv.org/content/early/2020/04/21/2020.04.12.20062794

35. Tomar, A., Gupta, N.: Prediction for the spread of covid-19 in india and effectiveness of preventive measures. Science of The Total Environment p. 138762 (2020)

36. Verma, M.K., Asad, A., Chatterjee, S.: Covid-19 pandemic: Power law spread and flattening of the curve. Transactions of the Indian National Academy of Engineering (2020). DOI 10.1007/s41403-020-00104-y. URL https://doi.org/10.1007/s41403-020-00104-y

37. Walker, P., Whittaker, C., Watson, O., Baguelin, M., Ainslie, K., Bhatia, S., Bhatt, S., Boonyasiri, A., Boyd, O., Cattarino, L., et al.: Report 12: The global impact of covid-19 and strategies for mitigation and suppression (2020)

38. World Health Organization Situation Report as on 11th August 2020: URL https://www.who.int/docs/default-source/coronaviruse/situation-reports

